# DeepProg: an ensemble of deep-learning and machine-learning models for prognosis prediction using multi-omics data

**DOI:** 10.1101/19010082

**Authors:** Olivier Poirion, Zheng Jing, Kumardeep Chaudhary, Sijia Huang, Lana X. Garmire

## Abstract

Multi-omics data are good resources for prognosis and survival prediction, however, these are difficult to integrate computationally. We introduce DeepProg, a novel ensemble framework of deep-learning and machine-learning approaches that robustly predicts patient survival subtypes using multi-omics data. It identifies two optimal survival subtypes in most cancers and yields significantly better risk-stratification than other multi-omics integration methods. DeepProg is highly predictive, exemplified by two liver cancer (C-index 0.73-0.80) and five breast cancer datasets (C-index 0.68-0.73). Pan-cancer analysis associates common genomic signatures in poor survival subtypes with extracellular matrix modeling, immune deregulation, and mitosis processes. DeepProg is freely available at https://github.com/lanagarmire/DeepProg

## Background

Most survival-based molecular signatures are based on one single type of omics data ^1^. Since each omic platform has specific limitations and noises, multi-omics based integrative approach presumably can yield more coherent signatures ^2^. However, this approach to predict clinical phenotypes is much less explored comparatively, due to the combination of computational and practical challenges. These challenges include platform-specific measurement biases ^3^, different data distributions which require proper normalizations ^4^, as well as very limited sample-sizes with multi-omics measurements due to the high cost^5^.

Among the multi-omics data integration methods, most of them do not model patient survival as the objective, rather the survival differences associated with molecular subtypes are evaluated in a *post hoc* fashion ^6^. Moreover, many methods use unsupervised approaches, unsuitable for predicting new patient statuses iCluster, Similarity Network Fusion (SNF), MAUI, and Multi-Omics Factor Analysis (MOFA+) are such examples^7–10^. iCluster is of the earliest methods to cluster cancer samples into different molecular subtypes based on multi-omic features, using probabilistic modelling to project the data to a lower embedding ^7^. Similarity Network Fusion (SNF) algorithm is another popular clustering method to integrate different omic features, by first constructing a distinct similarity network for each omic then fusing the networks using an iterative procedure ^8^. It was applied on multiple TCGA cancer datasets ^11,12^. MAUI is a non-linear dimension reduction framework for multi-omic integration which uses variational autoencoder to produce latent features that can be used for either clustering or classification ^9^. Similarly, MOFA+ is a statistical framework using factor analysis through standard matrix factorization to infer latent variables from multi-omic datasets explaining the most source of variation ^10^.

Identifying disease subtypes is clinically very significant. For example, it is generally accepted that cancer of a specific organ has multiple subtypes. Using molecular signatures to identify cancer subtypes allows tumor classification beyond tumor stage, grade, or tissue of origin^13^. Cancer subtypes sharing similar molecular and pathway alterations could be treated with the same drugs ^14^. One of such subtypes is survival stratified patient subtypes based on prognostic signatures ^15^. Once inferred, their signatures can be used as starting points for follow-up therapeutic or prognostic studies ^16^. Moreover, the molecular differences associated with patient survival help shed light to understand the mechanism of tumor progression ^17^. The gained knowledge not only helps to improve disease monitoring and management, but also provides information for prevention and treatment.

Here we propose a unique computational modeling framework called DeepProg, different from all methods mentioned above. It explicitly models patient survival as the objective and is predictive of new patient survival risks. DeepProg constructs a flexible ensemble of hybrid-models (a combination of deep-learning and machine learning models) and integrates their outputs following the ensemble learning paradigm. We applied DeepProg on RNA-Seq, Methylation, and miRNA data from 32 cancers in The Cancer Genome Atlas (TCGA), with a total of around 10,000 samples. DeepProg shows better predictive accuracies when compared to SNF based multi-omics integration method and the baseline Cox-PH method. The gene expression in the worst survival subtype of all cancers shares common signatures involved in biological functions such as mitotic enhancement, extracellular-matrix destabilization, or immune deregulation. Moreover, DeepProg can successfully predict the outcomes for samples from one cancer using the models built upon other cancers. In short, DeepProg is a powerful, generic, machine-learning and deep-learning based method that can be used to predict the survival subtype of an individual patient.

## Methods

### TCGA datasets

We obtained the 32 cancer multi-omic datasets from NCBI using TCGA portal (https://tcga-data.nci.nih.gov/tcga/). We used the package TCGA-Assembler (versions 2.0.5) and wrote custom scripts to download RNA-Seq (UNC IlluminaHiSeq RNASeqV2), miRNA Sequencing (BCGSC IlluminaHiSeq, Level 3), and DNA methylation (JHU-USC HumanMethylation450) data from the TCGA website on November 4-14^th^, 2017. We also obtained the survival information from the portal: https://portal.gdc.cancer.gov/. We used the same preprocessing steps as detailed in our previous study ^18^. We first downloaded RNA-Seq, miRNA-Seq and methylation data using the functions *DownloadRNASeqData, DownloadmiRNASeqData*, and *DownloadMethylationData* from TCGA-Assembler, respectively. Then, we processed the data with the functions *ProcessRNASeqData, ProcessmiRNASeqData*, and *ProcessMethylation450Data*. In addition, we processed the methylation data with the function *CalculateSingleValueMethylationData*. Finally, for each omic data type, we created a gene-by-sample data matrix in the Tabular Separated Value (TSV) format using a custom script.

### Validation datasets

For breast cancer data, we use four public breast cancer gene expression microarray datasets and one Metabric RNA-Seq dataset as the validation datasets. Four public datasets (all on Affymetrix HG-U133A microarray platform) were downloaded from Gene Expression Omnibus (GEO). Their accession IDs are GSE4922 ^19^, GSE1456 ^20^, GSE3494 ^21^ and GSE7390 ^22^. Their pre-processing was described in a previous study ^23^. For the Metabric dataset, we obtained approval from the Synapse repository: https://www.synapse.org/#!Synapse:syn1688369, and used the provided normalized data described in the Breast Cancer Challenge ^24^. The metabric dataset consists of 1981 breast cancer samples, from which we extracted RNA-Seq data. For hepatocarcinoma datasets, we used two larger datasets: LIRI and GSE datasets, as described in the previous study ^18^.

### DeepProg framework

DeepProg is a semi-supervised flexible hybrid machine-learning framework that takes multiple omics data matrices and survival information as the input. For each sample *s*, the survival data have two features: the observation time *t* and the observed event (death) *e*. The pipeline is composed of the following unsupervised and supervised learning modules (the detail of each step is described in the subsequent paragraphs). Module 1: unsupervised subtype inference: each input matrix is processed with: a) normalization, b) transformation using an autoencoder for each omics data type, and c) selection of the survival-associated latent-space features from the bottle neck layer of autoencoders. The selected survival-associated latent-space features from all the omics are then combined for clustering analysis. Module 2: supervised prediction of a new sample, this module is composed of the following steps: a) construction of a classifier using the training set, b) selection and normalization of the common features with the new sample, c) prediction. For both unsupervised and supervised inferences, we use an ensemble of DeepProg models through boosting approach: each model is constructed with a random subset (80%) of the training dataset. The clustering and the prediction results are combined according to the relevance of each model.

### Normalization

As default, DeepProg first selects the top 100 features from the training set that have the highest variance. Then for each sample, we inversely rank the features and divide them by 100, so that the score is normalized between 0 and 1. Next we compute the sample-sample Pearson correlation matrix of size *n*, the number of samples. For each sample, we use the sample-sample distances as new features and normalize them using the rank as well. As a result, each sample has *n* features with the score of the first feature equal to 1.0 and the last feature equal to 0.

To normalize a new sample (in the model prediction stage), we first select the set of common features between the new sample and the training set. We then perform the same steps as described above: a) selection of top 100 features, b) rank-based normalization, c) distance computation with the samples from the training set, and d) rank normalization.

### Autoencoder transformation

An autoencoder is a function *f*(*v*) = *v*′’ that reconstructs the original input vector *v* composed of *m* features through multiple nonlinear transformations (*size*(*v*) = *size*(*v*′) = *m*). For each omic data type, we create one autoencoder with one hidden layer of size *h* (default 100) that corresponds to the following equation:

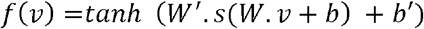

*W*’, *W* are two weight matrices of size *h by m* and *m by h*, and b, b’ are two bias vectors of size *h* and *h*’. *tanh* is a nonlinear, element-wise activation function defined as

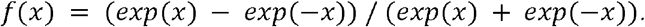

To train our autoencoders, we search the optimal *W*, W’*, b\** and *b*’*\** that minimizes the log-loss function.

We use python (2.7) Keras package (1.2.2) with theano as tensor library, to build our autoencoders. We use the Adam optimization algorithm to identify *W*, W’*, b\** and *b*’*\**. We train our autoencoder on 10 epochs and introduce 50% of dropout (i.e. 50% of the coefficients from W and W’ will be randomly set to 0) at each training iteration.

### Hyperparameter tuning

To help selecting the best set of hyperparameters (i.e. number of epochs, network shape, dropout rate…), DeepProg has an optional hyperparameter tuning module based on Gaussian optimization and it relies on the *scikit-optimize* (https://scikit-optimize.github.io/stable/) and the *tune* (https://docs.ray.io/en/latest/tune.html) python libraries. The computation of the ensemble of models and/or the hyperparameters grid-search can optionally be distributed on multiple nodes and external supercomputers using the python ray framework (https://docs.ray.io/en/latest/).

### Selection of new hidden-layer features linked to survival

For each of the transformed feature in the hidden layer, we build a univariate Cox-PH model using the python package *lifelines* (https://github.com/CamDavidsonPilon/lifelines) and identify those with log-rank p-values (Wilcoxon test) < 0.01. We then extract all the significant new latent features from all autoencoders and combine them as a new matrix Z.

### Cancer subtype detection

The default clustering method to identify subtypes is the gaussian mixture model-based clustering. We use the *GaussianMixture* function from the scikit-learn package with 1000 iterations, 100 initiations and a diagonal covariance matrix. The resulting clusters are sorted according to their median survival levels: the cluster labelled as “0” has the overall lowest median survival, while the last cluster “N” has the highest survival overall. Other clustering methods, K-means and dichotomized Lasso Cox-PH model, can replace the default gaussian mixture method.

### Construction of supervised classifiers to predict the cancer subtype in new samples

We use the cluster labels obtained from the above Gaussian mixture model to build several supervised machine learning models that can classify any new sample, under the condition that they have at least a subset of features in common with those input features from the training set. First, we compute a Kruskal-Wallis test for each omic type and each feature, in order to detect the most discriminative features with respect to the cluster labels. Then we select the 50 most discriminative features for each omic type and combine them to form a new training matrix M. We apply Support Vector Machine (SVM) algorithm to construct a predictive model using M as the input and the cluster labels as classes. To find the best hyper-parameters of the classifier, we perform a grid-search using a 5-fold cross-validation on M, with the objective to minimize the errors of the test fold. The algorithm constructs at first a classifier using all the omic types from the training samples. If a new sample shares only a subset of omics data types and a subset of the features with the training samples (eg. a sample has only RNA-Seq measurement), then DeepProg constructs a classifier using only this subset of omics data type and features, before applying it to the new sample. We use the python *sklearn* package to construct SVM models and infer the class probability with the *predict_proba* function, by fitting a logistic regression model on the SVM scores ^25^.

### Boosting procedure to enhance the robustness of DeepProg

To obtain a more robust model, we aggregate multiple DeepProg models constructed on a random subset of the training samples. As the default, we use 10 models with 80% of original training samples to construct all the cancer models, except for LUSC and PRAD which we use 20 models since they are more difficult to train. The aggregation of these models (per cancer) works as the following: after fitting, we eliminate those models without any new features linked to survival or having no cluster labels significantly associated with survival (log-rank p-value > 0.05). For a given sample, the probability of belonging to a particular cancer subtype is the average of the probabilities given by all the remaining models. We use the class probability of the worst survival subtype to assign the final label.

### Choosing the correct input number of clusters and performance metrics

When fitting a model, DeepProg computes several quality metrics: the log-rank p-value for a Cox-PH model using the cluster labels from all or only the hold-out samples as described above, the concordance index (C-index) ^26^, and the Silhouette score measuring the clusters homogeneity. In addition, DeepProg measures the clustering stability, that is, the consistency of class labeling among the different models during boosting. We compute the clustering as the following: *a)* For each pair of models, we compute the adjusted Rand Index between the two set of cluster labels (ARI) ^27^, *b)* we then calculate the mean of all the pair-wise rand indexes. For each cancer model we test different initial number of clusters (K=2,3,4,5). We then select the K presenting the best overall results based on silhouette score. Furthermore, we also select carefully the K that minimizes the crossovers on the Kaplan-Meier (KM) plots, when plotting the stratified patient survival groups according to the cluster labels.

To identify the input omics features differentially expressed between the worst survival subtype and other(s), we perform two-group (worst survival subgroup and the other remaining samples) Wilcoxon rank-sum test for each feature, using the Scipy.stats package. We then select features significantly over- or under-expressed with p-values<0.001. Next, we rank the differentially expressed features among the 32 cancers. For this purpose, we construct a Cox-PH model for each cancer and each significant feature and rank the features according to their -log10 (log-rank p-value). We then normalize the ranks among these significant features between 0 and 1, where 1 is attributed to the feature with the lowest Cox-PH log-rank p-value and 0 is assigned to the feature with the highest Cox-PH log-rank p-value in the set. We then sum the ranks of each feature among the 32 cancers to obtain its final score.

### Impact of tumor heterogeneity for BRCA and HCC on DA genes

We use xCell web interface ^28^ to infer the tumor composition amongst 67 reference cell types. We construct l1-penalized logistic regression using the *statsmodels* python library model, *fit_regularized* function from the *logit* class with alpha=1.0 for each gene with and without the tissue composition as cofounders and using the cluster labels as outcome. Prior to the regression, we scale the features to have mean=0 and std=1 for each features using the *RobustScaler* from scikit-learn. We rank the significant (the two-tailed t-stats p-values <0.05) coefficient for the two types of models and compared their overall similarities using the Kendall-Tau correlation measurement similar to before ^29^.

### Comparison with other data integration methods

To infer clusters from SNF, we use rpy2 to call SNF from python with the ‘*ExecuteSNF*’ function from the *CancerSubtypes* R library (v1.16) with the default parameters and use the same number of clusters (*k*) for DeepProg. We also substitute the autoencoder step of the DeepProg configuration with two other matrix factorization methods: MOFA+ and MAUI, using TCGA HCC and BRCA datasets. In each alternative approach, we transform the multi-omic matrices into 100 new components, followed by the same remaining steps in DeepProg (eg. survival associated feature filtering, clustering). For MOFA+ method (package *MOFA*), we obtain 100 features using the following parameters *iterations=500, convergence_mode=‘slow*’, *startELBO=1, freqELBO=1*. For MAUI (package *maui* for python3, Released: Sep 17, 2020), we obtain 100 latent features using the following parameters *learning rate = 0*.*0001, epochs=500, one hidden layer of 1100 nodes*. However, none of the 100 features is significantly (P<0.05) associated with survival in Cox-PH regression step of DeepProg workflow. Finally, we also substitute the autoencoder step by standard PCA using the *scikit*-*learn* python library.

### Construction of the co-expression network

For each cancer, we first identify (at most) the top 1000 RNA-Seq genes enriched in the worst subtype according to their Wilcoxon rank test p-value. We then use these genes to construct a Gene Regulatory Network. For each pair of genes (nodes), we obtain an interaction score based on their correlations, and assign it to the edge between them. For each network, we normalize the scores by dividing them with the maximal value. We then score each gene with the sum of its edge scores. We combine the network obtained for the 32 cancers into a global pan-cancer co-expression network, using the GRNBoost2 algorithm from the python package *arboreto*. Specifically, we use the following steps: *a)* aggregating the nodes, node weights, edges, and edge weights of each cancer network into a consensus graph, *b)* selecting the top 200 genes and construct its corresponding subgraph, *c)* performing edge pruning on each gene by removing all but the top 10 edges, according to their weights, and *d)*applying a community detection algorithm on the graph using the random-walk algorithm from the python library igraph, and visualizing the graph using Gephi ^30^.

### Code availability

The source code and documentation for the DeepProg framework is free for non-commercial use under GPL v3 license at: https://github.com/lanagarmire/DeepProg. The workflow is written in Python3 and was tested under Linux, OSX, and Windows. The package contains instructions for installation and usage, and the different requirements. A docker image containing all the dependencies installed is also freely available at: https://hub.docker.com/repository/docker/opoirion/deepprog_docker.

## Results

### DeepProg Method overview

DeepProg is a general hybrid and flexible computational framework to predict patient survival based on one or more omics data types, such as mRNA transcriptomics, DNA methylation and microRNA expression (**Figure 1)**. The first phase of DeepProg is composed of dimension reduction and feature transformation using custom rank normalizations and auto-encoders, a type of deep neural-network. It uses “modularized” design for auto-encoders, where each data type is modeled by one auto-encoder, to allow flexibility and extendibility to heterogeneous data types. In the default implementation, the auto-encoders have 3 layers, the input layer, the hidden layer (100 nodes), and the output layer. The transformed features are then subject to uni-variate Cox-PH fitting, in order to further select a subset of features linked to survival. Next, using unsupervised clustering approach, DeepProg identifies the optimal number of classes (labels) of survival subpopulations, and uses these classes to construct support vector machine (SVM) based machine-learning models, in order to predict a new patient’s survival group. To ensure the robustness of the models, DeepProg adopts a boosting approach and builds an ensemble of models. The boosting approach yields more accurate p-values and C-indices with lower variances and leads to faster convergence of the models (**Additional File 1: Table S1**). Each of these models is constructed with a random subset (eg. 4/5) of the original dataset and evaluated using the C-index value from the remaining hold-out (eg. 1/5) testing samples. For efficiency, the computation of DeepProg is fully distributed, since each model can be fit separately.

**Figure 1.**
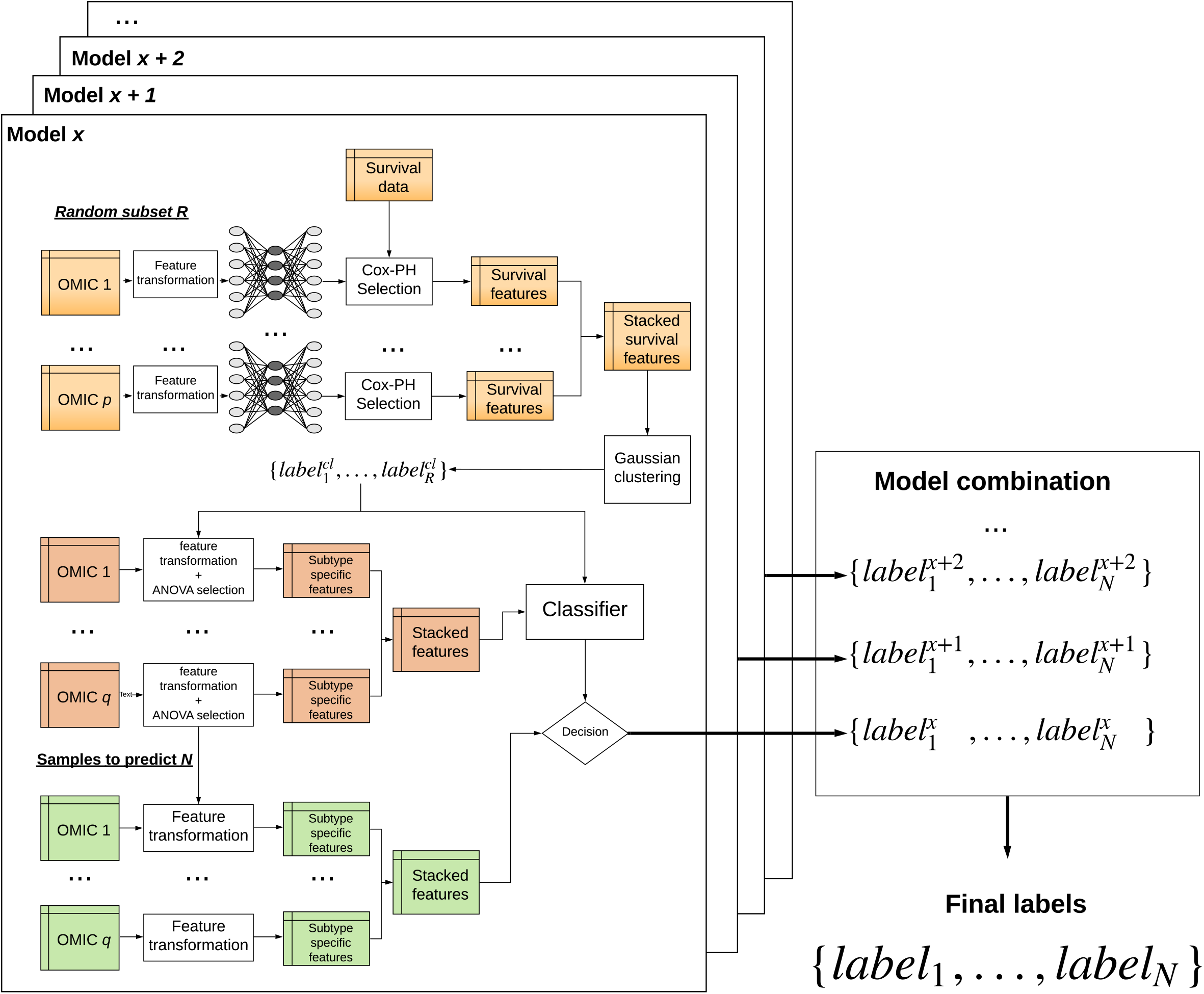
The computational framework of DeepProg. DeepProg uses the boosting strategy to build several models from a random subset of the dataset. For each model, each omic data matrix is normalized and then transformed using an autoencoder. Each of the new hidden-layer features in autoencoder is then tested for association with survival using univariate Cox-PH models. The features significantly associated with survival are then subject to clustering (Gaussian clustering by default). Upon determining the optimal cluster, the top features in each omic input data type are selected through Kruskal-Wallis analysis (default threshold = 0.05). Finally, these top omics features are used to construct a support vector machine (SVM) classifier, and to predict the survival risk group of a new sample. DeepProg combines the outputs of all the classifier models to produce more robust results.

### Prognostic prediction on 32 TCGA cancers

We applied DeepProg to analyze the multi-omics data (RNA-Seq, miRNA-Seq and DNA methylation) of 32 cancers in TCGA **(Additional File 2: Table S2)**. We used only RNA and MIR for Ovarian Cancer (OV) because only a small fraction (9 out of 300) of the samples had the 3-omics data at the time of the manuscript submission. For each cancer type, we selected the optimal clustering number K that produces the best combination of silhouette scores and Adjusted Rand Index (**Additional File 3: Table S3**), metrics that measure the clustering stabilities and accuracy. Almost all cancers (30 out of 32) have K=2 as the most optimum survival-subgroups (**Figure 2A**). With the optimal cluster numbers, we computed the log-rank p-values among the different survival subtypes of each cancer, all of which are statistically significant (log-rank p-values < 0.05) and have C-indexes (0.6 -1.0) greater than 0.5, the expected value of random models. Among them, 23 out of 32 cancers have log-rank p-values less than 5e-4, highlighting the values of the models at differentiating patient survival (**Figure 2B**). Additionally, we investigated the average number of hidden-layer features significantly associated with survival, for each omic data type and each cancer (**Additional File 4: Fig. S1**). Overall, RNA-Seq has the most amount of important hidden features towards survival prediction. miRNA hidden features have similar patterns in all cancers, with fewer total counts. Although vast heterogeneity exists among 32 cancers, some cancers known to be closely related, such as colon cancer (COAD) and gastric cancer (STAD), as well as bladder cancer (BLCA) and kidney cancer (KIRC), also share similar prognostic hidden features.

**Figure 2.**
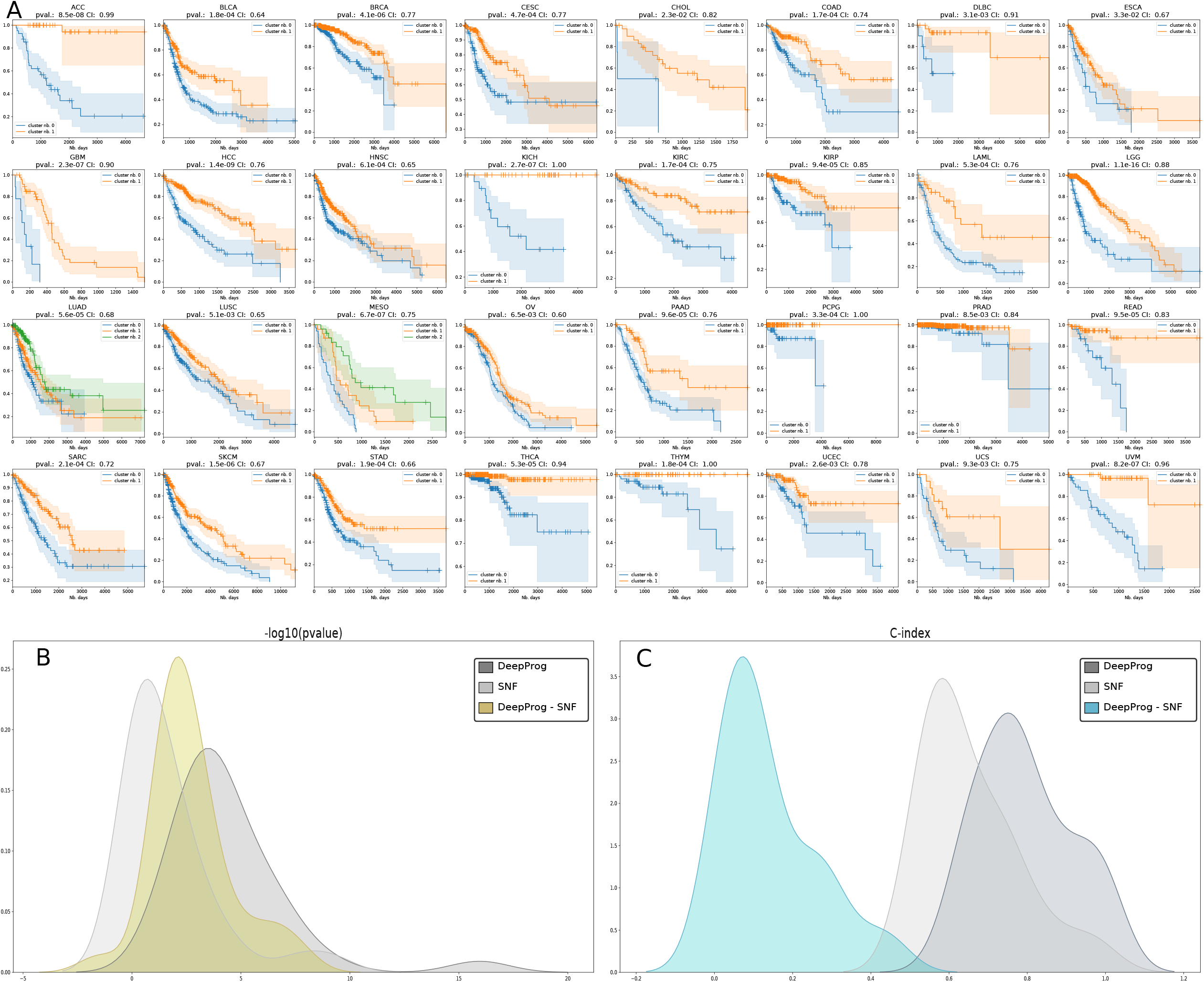
DeepProg performance for the 32 TCGA cancer datasets. A) Kaplan-Meier plots for each cancer type, where the survival risk group stratification is determined by DeepProg. B) The density distributions of -log10 (log-rank p-value) for the Cox-PH models based on the subtypes determined by DeepProg (light grey line), SNF (dark grey line), or the pair-wise -log10 (log-rank p-value) differences between DeepProg and SNF (blue line). C) Smoothed C-index distributions for the Cox-PH models based on the subtypes determined by DeepProg (light grey line), SNF (dark grey line), or the pair-wise C-index difference between DeepProg and SNF (blue line).

We previously showed that for the prototype of DeepProg, adding clinical variables such as cancer stage, ethnicities, etc did not help improving the predictive results in HCC ^18^. Here we also directly compare the performance of DeepProg vs. a simple model based on stage stratification (stage I+II vs. Stage III+IV) on 32 cancer types. As shown in **Additional File 4: Fig. S2A**, DeepProg has significantly better (Rank sum test p-value=2.4e-3) log-rank p-values compared to the simple model based on stage-stratified survival difference. To further demonstrate the DeepProg is capable to predict patient survival outcome beyond tumor stage, we next focused on late stage (Stage III and IV) COAD and STAD. We constructed DeepProg models using patients from the latter stages III and IV and compared their results to survival difference using stage III vs IV for stratification (**Additional File 4: Fig. S2B-E**). The subtypes for STAD and COAD identified by DeepProg are clearly more significant (log-rank p-values of 5.5e-04 and 2.7e-06 respectively) than those based on tumor stages (log-rank p-values of 0.16 and 0.012 respectively). Moreover, the subtypes from DeepPorg are not significantly associated with the stage (Fisher exact p-values of 0.08 and 0.14 for COAD and STAD, respectively). Thus, DeepProg provides much more information to predict patient survival than the clinical factor tumor stage.

### Comparison between DeepProg and other methods

To evaluate the new DeepProg method, we compared the results from the 32 cancers above with those obtained from the Similarity Network Fusion (SNF) algorithm ^8^, a state-of-the-art method to integrate multi-omics data (**Figure 2B and 2C, Additional File 4: Fig. S3**). Previously SNF was used to identify cancer subtypes linked to survival by others ^11,12^. As shown in **Figure 2B**, the survival subtypes from SNF only have significant survival difference in 13 out of 32 cancers (p-value<0.05). In all, DeepProg yields much better log-rank p-values (**Figure 2B**) and C-indices (**Figure 2C**). Additionally, considering that TCGA datasets might have changed since the time of the SNF publication, we also used the patient subtypes identified in the original SNF paper on five test datasets, and used them to obtain log-rank survival subtype p-values ^8^. These p-values are all less significant, compared to those obtained from DeepProg using the same five datasets as the inputs (**Additional File 5: Table S4**).

We also substituted the autoencoder step of the DeepProg configuration with a simple PCA decomposition and two matrix factorization methods including MAUI and MOFA+, using TCGA HCC and BRCA datasets. In each alternative approach, we transformed the multi-omic matrices into 100 new components, followed by the same remaining steps in DeepProg (eg. survival associated feature filtering, clustering). While none of the 100 features from MAUI has a p-value <0.05 in Cox-PH filtering step, PCA and MOFA+ both have much worse performances compared to the default dimension reduction step in DeepProg (**Figure 3 and Additional File 6: Table S5**). On HCC testing data, C-indices from PCA and MOFA+ are 0.60 and 0.59 respectively **(Figure 3A-B**), compared to that of 0.76 by DeepProg (**Figure 3C**). On BRCA testing data, PCA and MOFA+ yield C-indices of 0.58 and 0.62 respectively (**Figure 3D-E**), whereas DeepProg has a C-index of 0.77 (**Figure 3F**). In conclusion, based on the HCC and BRCA benchmarks, DeepProg is significantly better to infer subtypes linked to survival from multi-omics data compared to the matrix factorization methods and PCA.

**Figure 3.**
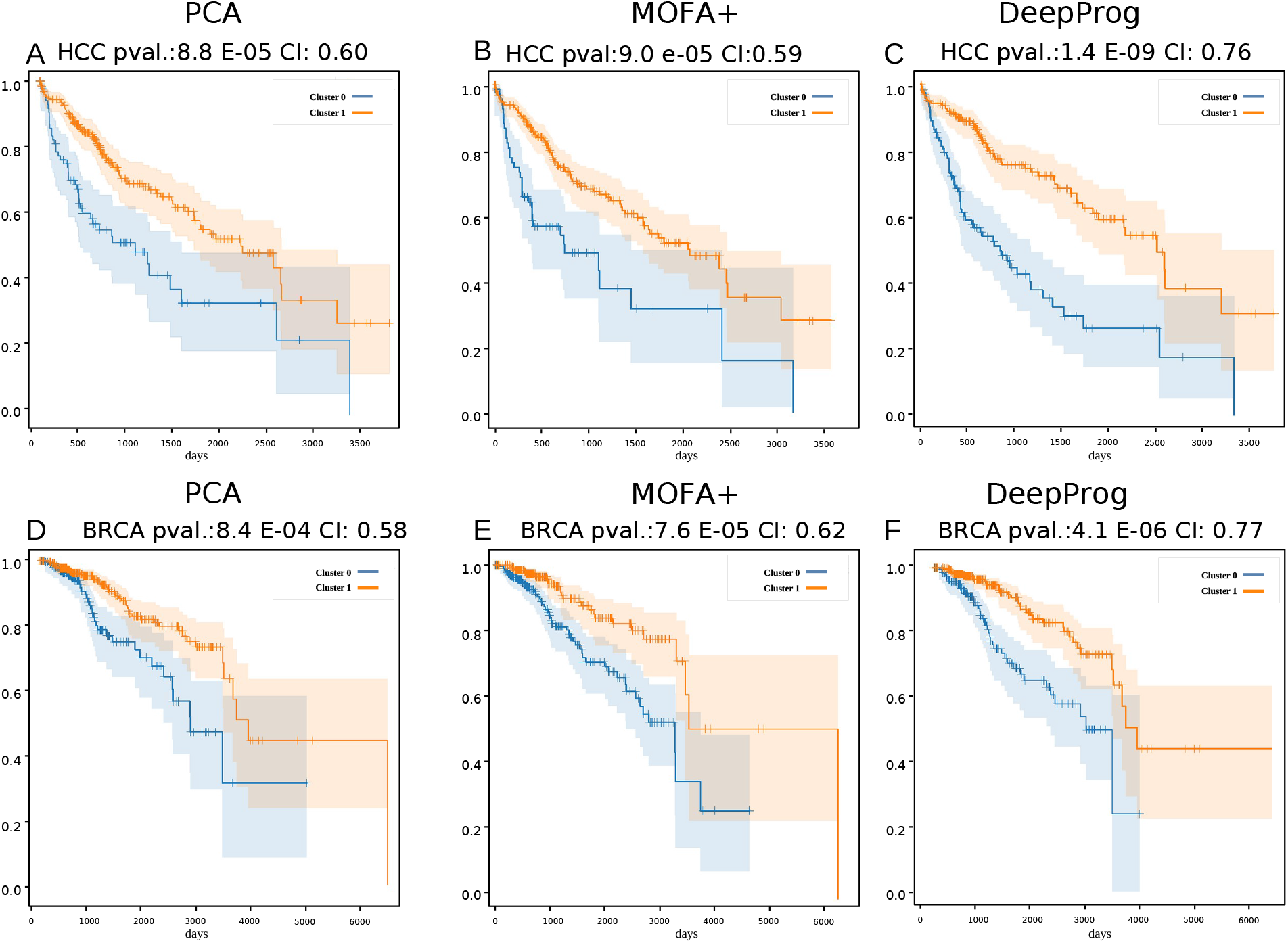
Comparing the performance of DeepProg and its variations, where the default autoencoder is substituted by a simple PCA decomposition or MOFA+ method to generate an input matrix of the same dimensions, using TCGA HCC (A-C) and BRCA (D-F) datasets. Methods in comparison: (A, D): PCA; (B, E): MOFA+; (C, F): DeepProg default.

Lastly, we compared DeepProg with a baseline model, where the Z-score normalized features are directly fit by the Cox-PH model with Lasso penalization without the autoencoder step. The samples are dichotomized into the same number of clusters as in DeepProg, subjective to the same parameterization whenever applicable. On the same hold-out samples across 32 cancers, DeepProg shows significantly better log-rank p-values (p-value<0.0005, 2-sided t-test) than the baseline Cox-PH model (**Additional File 4: Fig. S4**).

### Validation of DeepProg performance by other cohorts

One key advantage of the DeepProg workflow is its ability to predict the survival subtype of any new individual sample that has some common RNA, miRNA or DNA methylation features with the training dataset (**Figure 1B)**. DeepProg normalizes a new sample by taking the relative rank of the features and use them to compute the distances to the samples in the training set (see **Methods**). To validate the patient survival risk stratification of DeepProg models, we applied them on additional independent cancer datasets, two from hepatocellular carcinoma (HCC) cohorts (**Figure 4A-B**) and four from breast cancer (BRCA) cohorts (**Figure 4C-F**). The two HCC validation sets are LIRI dataset with 230 RNA-Seq samples, and GSE dataset with 221 gene expression array results (see **Methods**). We obtained a C-index of 0.80 and log-rank p-value of 1.2e-4 (LIRI), and a C-index of 0.73 and log-rank p-value of 1.5e-5 (GSE), respectively (**Figure 4A-B**). The four BRCA datasets have C-indices of 0.68-0.73, all with significant log-rank p-values (<0.05) for survival difference (**Figure 4C-F**). We thus validated the predictability of DeepProg by additional HCC and BRCA cohorts.

**Figure 4.**
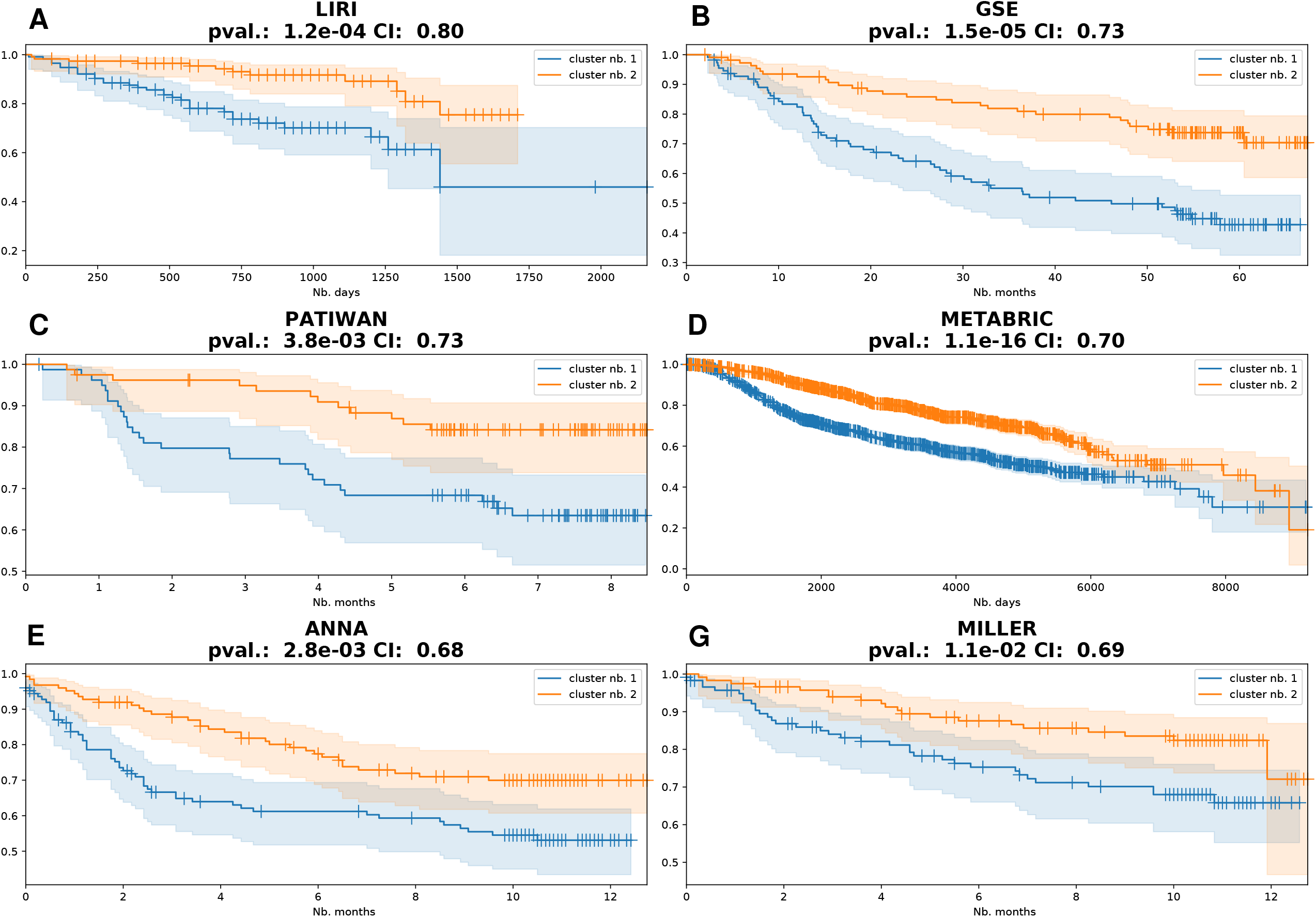
Validation of DeepProg subtype predictions by independent breast cancer and liver cancer cohorts. RNA-Seq Validation datasets for HCC: (A) LIRI (n=230) and (B) GSE (n=221) and validation datasets for BRCA: (C) Patiwan (n=159), (D) Metabric (n=1981), (E) Anna (n=249), and (F) Miller (n=236).

### Identification of signature genes for the worst survival subtypes reveals pan-cancer patterns

In order to identify the key features that are associated with patient survival differences, we conducted a comprehensive analysis of features in each omic layer that are significantly over- or under-expressed among the subset of patients with the poorest survival. Next, among the over- or under-expressed features we selected important features from the input data types whose Wilcoxon rank test p-values are less than 1e-4. For each of these features we computed the univariate Cox-PH regression in each cancer type and ranked them based on the -log10 (p-values). Upon normalizing these ranks between 0 and 1, we obtained a pan-cancer rank by summing over all 32 cancer types (see **Methods**). We describe the results in RNA-Seq analysis in the following and summarize the results on microRNA and DNA methylation analysis in **Additional File 7**.

The RNA-Seq analysis shows some emerging patterns of over-represented genes within the poorest survival group (**Figure 5A**). CDC20 is ranked first, and some other genes from the cell division cycle (CDC) family, including CDCA8, CDCA5, CDC25C and CDCA2, are also among the top 100 genes (**Additional File 8: Table S6**). Additionally, numerous genes from the Kinesin Family Member (KIF) (i.e. KIF4A, KIF2C, KIF23, KIF20A, KIF18A, KIFC1, KIF18B, and KIF14) are present in the top 100 genes (**Additional File 8: Table S6)**. The CDC genes ^31–35^ and KIF genes over expression ^36,37^ have been reported in the metastasis process and linked to poor prognosis. Many other genes over-expressed in the poor survival group are concordant with previous studies, such as ITGA5, CALU, PLKA1, KPNA2, APCDDL1, LGALS1, GLT25D1, CKAP4, IGF2BP3 and ANXA5 ^38^. Using the ranking values, we clustered the cancers and the genes and detected two clear gene clusters, enriched with biological functions of cell-cycle and mitosis (Adj. p-value= 3e-42) and Extracellular matrix organization pathway (Adj. p-value=6e-9), respectively (**Figure 5A**). In addition, the analysis shows two distinct groups of cancers, where GBM, HNSC, OV, STAD, COAD, LUSC, and KIRC are in one group, and cancers such as PRAD, PAAD, and LUAD are in the other group (**Figure 5A**).

**Figure 5.**
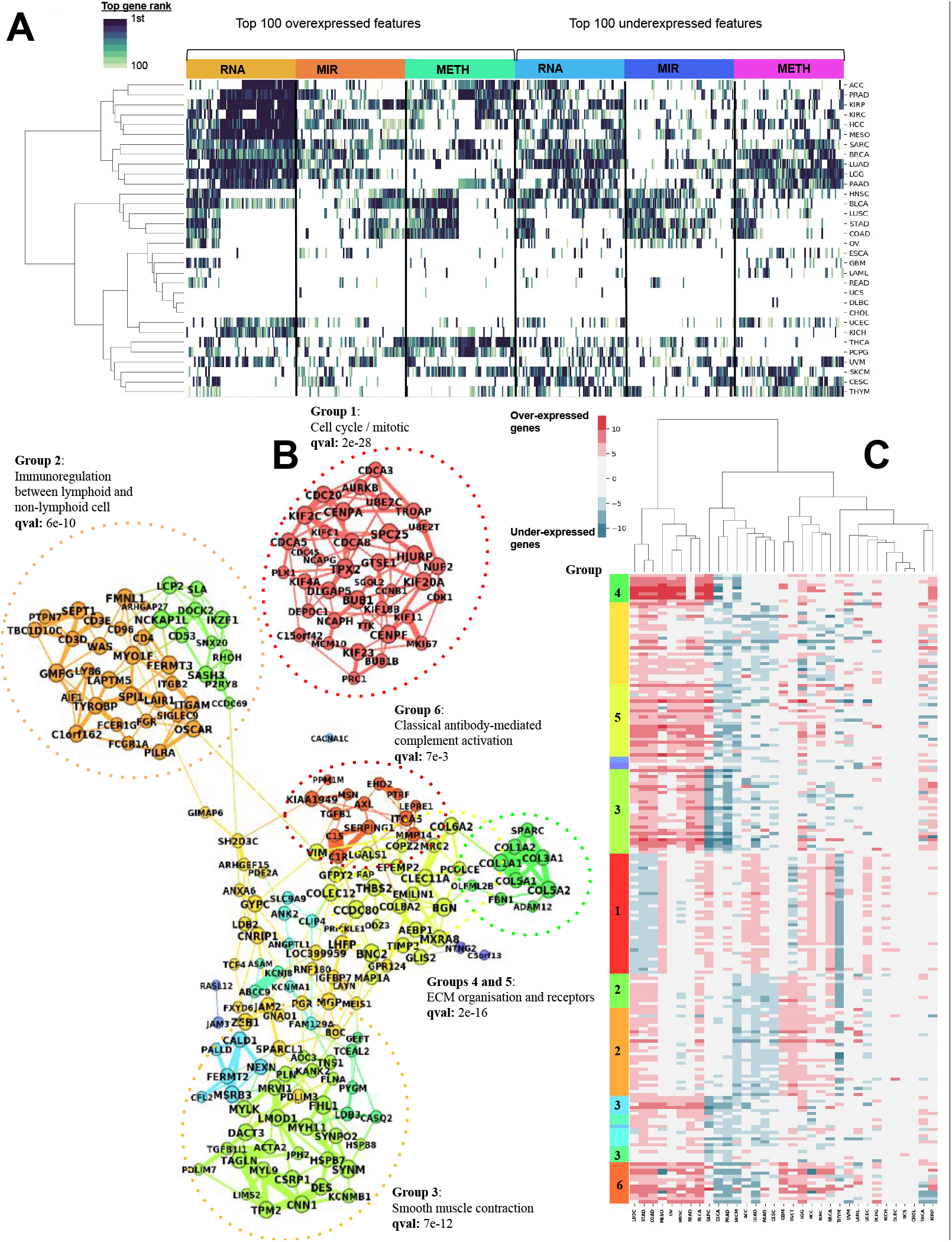
Pan-cancer analysis of RNA-Seq gene signatures in the worst survival vs. other groups. A) Top 100 over- and under-expressed genes for RNA, MIR, and METH omics ranked by survival predictive power. The colors correspond to the ranks of the genes based on their –log10 (log-rank p-value) of the uni-variate Cox-PH model. Based on these scores, the 32 cancers and the features are clustered using the WARD method. B) Co-expression network constructed with the top 200 differentially expressed genes from the 32 cancers. The 200 genes are clustered from the network topology with the Louvain algorithm. For each submodule, we identified the most significantly enriched pathway as shown on the figure. C) The expression values of these 200 genes used to construct the co-expression network. A clustering of the cancers using these features with the WARD method is represented in the *x-*axis

Among the genes that are under-expressed in the poorest survival groups, CBX7 and EZH1 are the top 2 genes (**Additional File 8: Table S6**). Down-regulation of CBX7 was shown to play a critical role in cancer progression ^39^. Similarly, EZH1 inhibition was shown to be involved in cell proliferation and carcinogenesis ^40,41^. Additionally, multiple genes in zinc finger family are downregulated (ZBTB7C, ZMAT1, ZNF18, ZNF540, ZNF589, ZNF554, and ZNF763). ZNF genes are a large family of transcription factor and many of them were shown relevance in cancer progression ^42^.

### RNA-Seq co-expression network analysis

To characterize further the RNA-Seq gene expression associated with the poorest survival subtypes, we performed a global gene co-expression analysis. For each cancer type, we selected differentially expressed genes from the worst survival subtype (**Figure 5A**) and constructed a pan-cancer consensus co-expression network. As an illustration, we constructed a subgraph of co-expression using the top 200 genes and the most significant edges (**Figure 5B**), performed gene community detection using random-walks algorithm ^43^. A large fraction of top co-expressed genes overlap with the top survival genes highlighted earlier. For example, a tight cluster (group 1) is composed of multiples CDC and KIF genes, together with BUB1, MCM10, AURKB, CENPA, CENPF, and PLK1. These genes are related to *mitosis and cell cycle pathway* (q-value=2e-28). Two clusters (groups 4 and 5) that include multiple collagens are enriched with *extracellular matrix (ECM) organization and receptors function* (q-value=2e-16) (**Figure 5B**). These results follow the conclusions of previous studies highlighting close correlations between ECM genes, notably SPARC and COL1A1, and tumor invasiveness ^44^. In addition, the network unveiled two major groups of genes associated with *immunoregulation between lymphoid and non-lymphoid cell* pathway (group 2, q-value=6e-10) and *smooth muscle contraction* (group 3, q-value=7e-12), respectively. Similar to signature gene results (**Figure 5A**), gene-cancer cluster map shows very close similarities between COAD and STAD on RNA co-expression (**Figure 5C**), which was observed earlier in features, and was reported as pan-gastrointestinal cancers from the cancer tissue-of-origin study (Hoadley et al., 2018).

To address potential confounding of tumor heterogeneity within patients, we used xCell ^28^ to deconvolute the cell types for each patient. We then adjusted the genes for all cell type compositions using logistic regression, similar to what we did before ^44^. We performed a comparison between before and post cell-type adjustment over the two sets of differentially expressed (DE) genes using Kendall-Tau correlation scores, similar to previous study ^29^. On HCC and BRCA, Kendall-Tau correlation scores are 0.52 (p-value < 1.04e-25) and 0.55 (p-value < 3.5e-150) respectively. The highly significant p-values reject the hypothesis that these two DE gene rankings are independent.

### Similar cancer types can be used as predictive models

Motivated by the similarities observed among some cancers, we explored if the models are suitable for transfer learning, that is, the model built on one particular cancer type can be used to predict survival of patients in another cancer type. We tested all pairs of 32 cancers, used alternatively as training and test datasets. Many of the cancer models are effective at predicting other cancer types (**Figure 6A**). Interestingly, models built on mesothelioma (MESO) data significantly predict the subtypes of 12 other cancer types, with long-rank p-values ranging from 0.048 to 4.8e-6, and C-indices ranging from 0.58 to 0.82. In general, cancer types that are biologically more relevant have higher predictive accuracies of cross-cancer predictions, for example, the cancer pair COAD/ STAD, the two close cancer types identified by the earlier signature gene analysis (**Figure 5C**). The STAD model significantly predicts the subtypes of COAD samples (p-value=0.018, CI=0.60) (**Figure 6B**), and vice versa for the COAD model prediction on STAD samples (p-value=5.4e-3, CI=0.66) (**Figure 6C**).

**Figure 6.**
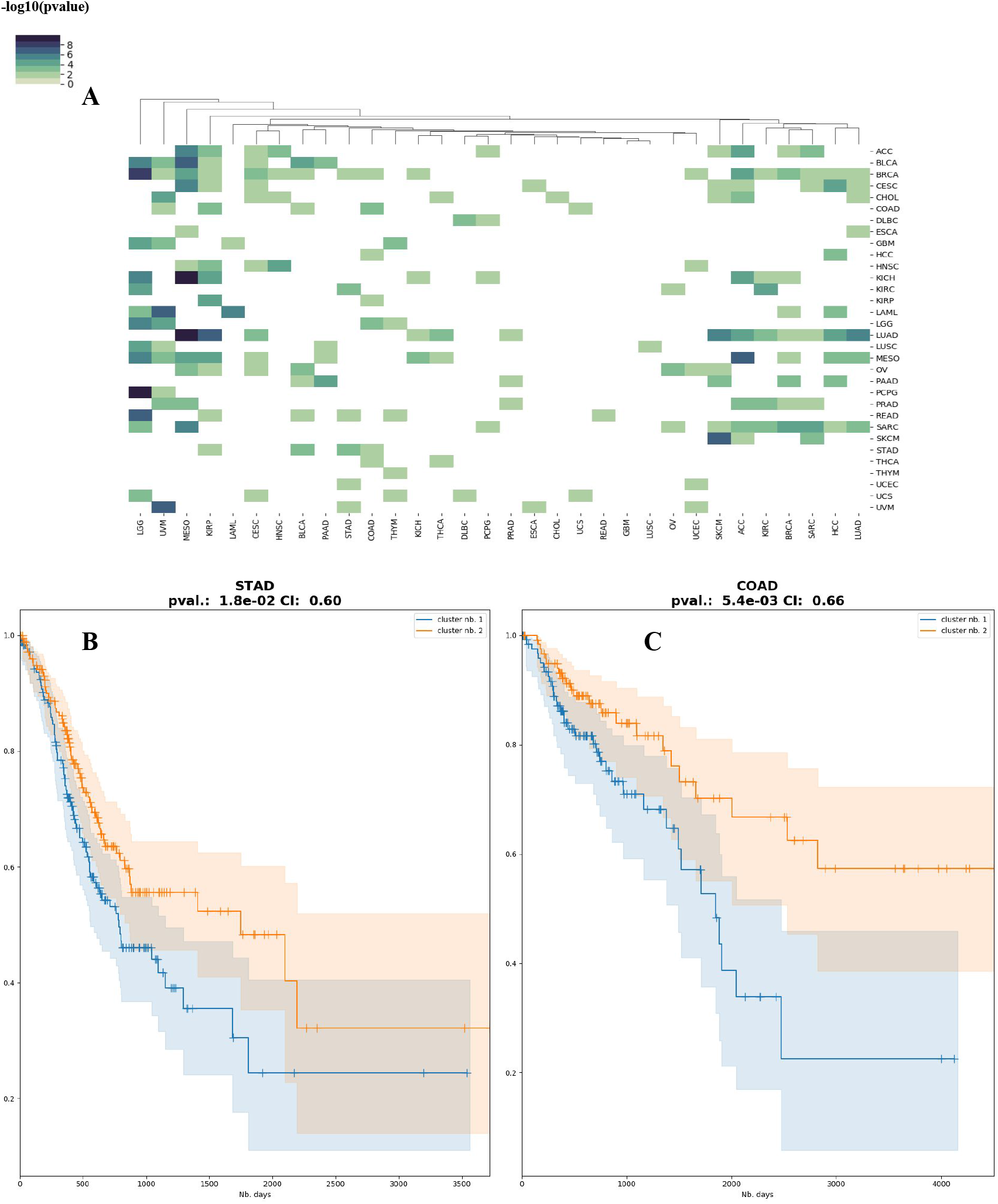
Transfer learning to predict survival subtypes of certain cancers using the DeepProg models trained by different cancers. A) Heatmap of the Cox-PH log-rank p-values for the subtypes inferred using each cancer as the training dataset. B) Kaplan-Meier plot of predicted subtypes for COAD, using the DeepProg model trained on STAD. C) Kaplan-Meier plot of predicted subtypes for STAD, using the DeepProg model trained on COAD.

Intrigued by the apparent lack of predictability between READ and COAD (**Figure 5A**), we investigated further the potential source. READ shows similar top 100 gene expression patterns with STAD and COAD, however, is quite different in the top 100 features at the miRNA and methylation levels (**Figure 5**). We thus constructed additional models for COAD and EAD using only RNA as features and were able to significantly improve the mutual predictabilities between COAD and READ (**Additional File 4 Fig. S5**).

## Discussion

In this report, we present a novel and generic computational framework, named DeepProg, which processes multiple types of omics data sets with a combination of deep-learning (autoencoder) and machine-learning algorithms, specifically for survival prediction. We have demonstrated several characteristics of DeepProg, including its superior predictive accuracy over the state-of-the-art method SNF, its robustness at predicting other HCC and BRCA population cohorts’ patient survival, as well as its suitability as a transfer learning tool trained from a relevant cancer to predict another cancer.

A few unique mechanistic features of DeepProg contribute to its accuracy. First, it uses boosting procedures that increase the robustness of the final model, by agglomerating weaker models from different subsets of the original samples. This design is well adapted to distributed computing architectures and can be scaled-up easily. Secondly, it employs a modularized design for each omic data type and can be extended to other omics and data types. In its default configuration DeepProg first processes each omic data set individually with autoencoders, and then merges the hidden layer features under a unified Cox-PH fitting – clustering – supervised classification workflow. The autoencoder structure transforms the initial input features of various omics types into new features. To our knowledge, DeepProg is the only multi-omic framework for survival subtyping. Another key aspect of DeepProg is the validation of the inferred labels using either internal *out-of-bag* samples and/or external datasets. In this study, we demonstrated the value of DeepProg in integrating 3 types of omics data: RNA-Seq, microRNA-Seq and DNA methylation. Other specialized deep-learning models to handle mutation or pathology image data can be developed as individual modules and added to DeepProg. DeepProg performs significantly better overall than SNF at predicting patient survival. One major reason is that SNF does not include survival information when performing integration, rather, it only relies on the patterns from multiple types of genomics data. Moreover, as an unsupervised method SNF cannot predict the prognosis of new sample(s) like DeepProg.

We further used DeepProg to identify global signatures of tumor aggressiveness among 32 types of cancers. Although previously several pan-cancer studies used one or different omic types to understand pan-cancer molecular hallmarks ^38,45,46^, the report here is the first of its kind to systematically characterize the differences between survival subtypes in pan-cancer. We identified the top survival features linked to the aggressive subtypes and focused on RNA-Seq expression analysis. The pan-cancer gene regulatory network highlights the top co-expressed genes in the most aggressive subtype of these cancers. Many of them are related to cell proliferation, extracellular matrix (ECM) organization, and immunoregulation, confirming earlier results in the literature on cancer invasion ^47,48^. Such genes are notably linked to the cell-division cycle ^33^, cytoskeleton structure ^49^, collagens^48^ or cadherin families ^50^. We also found several genes linked to smooth muscle contraction. For example, Calponin gene CNN1, TAGLN, and TMP2 are co-expressed in different cancers (Figure 4). Also, CNN1, TAGLN and TMP2 were already characterized as prognostic molecular markers for bladder cancer with higher expression associated with lower survival ^51^. Interestingly, various transcription factor families, such as Zinc finger genes are down-regulated and HOX genes are hypermethylated (**Additional File 8: Table S6**). Such observations are supported by previous reports, as multiple zinc-finger proteins have been shown to act as tumor suppressor genes ^52^, and dysregulation of HOX genes is frequent in cancer as many of them play important roles in cell differentiation ^53^. It will be of interest to follow up experimentally to test their effects. Lastly, through comprehensive comparison among 32 cancers, the molecular similarities that are clinically (survival) relevant are revealed. For example, aggressive subgroups from COAD and STAD, two gastroenteric cancers, present multiple common patterns. We speculate that these relationships can be exploited in the future to build more robust analyses ^54^, and help strategize treatment plans by leveraging patient profiles and cancer similarities.

## Conclusions

DeepProg is a novel ensemble framework of deep-learning and machine-learning approaches that robustly predicts patient survival subtypes using multi-omics data. We anticipate that DeepProg models are informative to predict patient survival risks, in diseases such as cancers.

## Data Availability

All multi-omics and survival data that support the findings of this study are available under figshare with the following identifier.

https://doi.org/10.6084/m9.figshare.14832813.v155

## Availability and requirements

**Project name:** DeepProg

**Project home page:** https://github.com/lanagarmire/DeepProg

**Operating system(s):** Linux, OSX, Windows, Docker

**Programming language:** Python, R

**Other requirements:** Python 3.8, Tensorflow, scikit-learn, scikit-survival, lifelines, Keras

**License:** PolyForm Perimeter License 1.0.0

**Any restrictions to use by non-academics:** license needed

## Declarations

### Ethics approval and consent to participate

All data utilized in this study has been previously published and is publicly available. The research performed in this study conformed to the principles of the Helsinki Declaration

### Consent for publication

Not Applicable

### Availability of data and materials

All multi-omics and survival data that support the findings of this study are available under “figshare” with the following identifier:

https://doi.org/10.6084/m9.figshare.14832813.v1^55^

## Competing interests

The authors declare that they have no competing interests

## Funding

This research was supported by grants K01ES025434 awarded by NIEHS through funds provided by the trans-NIH Big Data to Knowledge (BD2K) initiative (www.bd2k.nih.gov), P20 COBRE GM103457 awarded by NIH/NIGMS, R01 LM012373 and R01 LM012907 awarded by NLM, and R01 HD084633 awarded by NICHD to L.X. Garmire.

## Author Contributions

LG envisioned this project. OP developed DeepProg algorithm, implemented the project and conducted the analyses, OP and LG wrote the manuscript. ZJ helped analyzing the data, SH and KC helped to download and process the datasets. All authors have read and approved the final manuscript.

## Acknowledgements

We thank Dr. Bing He for testing the package and improving the documentation of the software.

## Supplementary materials

**Additional File 1 Table S1** DeepProg performances when using a number of models ranging from 1 to 30. The Cox-PH log-rank p-value (pval) and the C-index (CI) are calculated for two datasets: HCC and BRCA. Two validation datasets are used for HCC: LIRI and GSE, and four datasets for BRCA, named Anna, Patiwan, Miller, and Metabric datasets respectively.

**Additional File 2 Table S2** Summary of 32 TCGA cancer types.

**Additional File 3 Table S3** Cox-PH log-rank p-value, Silhouette score, and clustering stability score obtained using DeepProg on the 32 cancers and with a number of input clusters from 2 to 5.

**Additional File 4 (Figure S1-S5):** Additional information including details of DeepProg model and its comparison with other methods

**Figure S1**. Autoencoder hidden-layer features significantly associated with survival for each omic data type and cancer type.

**Figure S2**. Compare the performance of DeepProg vs. a simple model based on stage stratification (stage I+II vs. Stage III+IV) on 32 cancer types, on their distributions of the –log10(log-rank p-value) of stratified survival curves differences. And compare the performance of DeepProg vs. a simple model based on late stage (Stage III / IV) STAD (B-C) and COAD (D-E): DeepProg survival plots; (B, D): survival plots using stage stratification (C, E).

**Figure S3**. Kaplan-Meier plots for each cancer type, where the survival risk group stratification is determined by SNF, with the same datasets used from the DeepProg analysis in **Figure 2A**.

**Figure S4**. DeepProg has significantly better (more significant) log-rank p-values compared to the baseline Cox-PH model.

**Figure S5**. COAD and READ mutual predictability analysis using RNA as features. A) Subtypes inferred with DeepProg for COAD and used to predict READ subtypes. B) READ subtypes inferred with DeepProg and used to predict COAD subtypes.

**Additional File 5 Table S4** Comparison of DeepProg results using the same benchmark datasets as those obtained from the original SNF paper.

**Additional File 6 Table S5** Significances of survival-clusters inferred using MAUI, MOFA+, and PCA as alternatives to the autoencoder module in DeepProg, on TCGA HCC and BRCA datasets.

**Additional File 7**: Additional analysis of microRNA and methylation signatures between the worst survival subtype vs. the remaining samples in each cancer.

**Additional File 8 Table S6** Top 100 features significantly linked to the lowest survival subtypes among the 32 cancers and ranked using their individual Cox-PH p-value in the different datasets. The features are separated among RNA, MIR and METH features, by over- and under-expressed features.

## Notes

### Competing Interest Statement

The authors have declared no competing interest.

### Author Declarations

All relevant ethical guidelines have been followed and any necessary IRB and/or ethics committee approvals have been obtained.

Any clinical trials involved have been registered with an ICMJE-approved registry such as ClinicalTrials.gov and the trial ID is included in the manuscript.

